# TGF-β-pathway-based polygenic risk score modifies the association between red meat intake and colorectal cancer risk: Application of a novel pathway-based PRS method

**DOI:** 10.1101/2025.06.13.25329599

**Authors:** Joel Sanchez Mendez, Bryan Queme, Yubo Fu, John Morrison, Juan P. Lewinger, Eric Kawaguchi, Huaiyu Mi, Mireia Obón-Santacana, Ferran Moratalla-Navarro, Vicente Martín, Victor Moreno, Conghui Qu, Jeroen R Huyghe, Polly A Newcomb, Amanda I Phipps, Claire E Thomas, David V Conti, Jun Wang, Elizabeth A Platz, Kala Visvanathan, Temitope O Keku, Christina C. Newton, Caroline Y Um, Anshul Kundaje, Marc J Gunter, Niki Dimou, Nikos Papadimitriou, Franzel JB van Duijnhoven, Satu Männistö, Gad Rennert, Alicja Wolk, Michael Hoffmeister, Hermann Brenner, Yu Tian, Loïc Le Marchand, Emmanouil Bouras, Konstantinos K. Tsilidis, D Timothy Bishop, Robert Maclnnis, Daniel D Buchanan, Cornelia M. Ulrich, Anita R. Peoples, Andrew J Pellatt, Li Li, Matthew AM Devall, Demetrius Albanes, Sonja I Berndt, Stephen B Gruber, Edward Ruiz-Narvaez, Mingyang Song, David A Drew, Andrew T Chan, Marios Giannakis, Li Hsu, Ulrike Peters, Mariana C. Stern, W. James Gauderman

**Author notes:** **Correspondence**: W. James Gauderman, Division of Biostatistics, Department of Population and Public Health Sciences, University of Southern California, 1845 N Soto St, room SSB 202K, Los Angeles, CA 90032. Mariana C. Stern, University of Southern California, Keck School of Medicine, Norris Comprehensive Cancer Center, 1441 Eastlake Avenue, room 4455, Los Angeles, CA 90089.

## Abstract

**Background:** Red and/or processed meat are established colorectal cancer (CRC) risk factors. Genome-wide association studies (GWAS) have reported over 200 variants associated with CRC risk. We used functional annotation data to identify subsets of variants within known pathways to construct pathway-based Polygenic Risk Scores (pPRS) to assess interactions with meat intake.

**Methods:** A pooled sample of 30,812 cases and 40,504 CRC controls from 27 studies were analyzed. Quantiles for red and processed meat intake were constructed. 204 GWAS variants were annotated to genes with AnnoQ and assessed for overrepresentation in PANTHER-reported pathways. pPRS’s were constructed from significantly overrepresented pathways. Covariate-adjusted logistic regression models evaluated interactions between pPRS and red or processed meat intake in relation to CRC risk.

**Results:** A total of 30 variants were overrepresented in four pathways: Presenilin-Alzheimer disease, Cadherin/WNT-signaling, Gonadotropin-releasing hormone receptor, and TGF-β signaling. We found a significant interaction between TGF-β-pPRS and red meat intake (OR_int_ = 0.95; 95% CI = 0.92-0.98; p = 0.003). When variants in the TGF-β pathway were assessed, we observed significant interactions of red meat with rs2337113 (intron *SMAD7* gene, Chr18), and rs2208603 (intergenic region *BMP5*, Chr6) (p = 0.0005 & 0.036, respectively). There was no evidence of pPRS x red meat interactions for other pathways or with processed meat

**Conclusions:** This pathway-based interaction analysis revealed a statistically significant interaction between variants in the TGF-β pathway and red meat consumption that impacts CRC risk.

**Impact:** These findings shed light into the possible mechanistic link between red meat consumption and CRC risk.

**Impact statement:** In this work, we developed pathway-based Polygenic Risk Scores which, for the first time, suggested that red meat intake interacts with variants overrepresented in TGF-β signaling pathway to impact colorectal cancer risk.

## INTRODUCTION

Colorectal cancer (CRC) has the third highest cancer incidence, and the second highest cancer mortality worldwide (1). It is estimated that if current trends are maintained, 3.2 million new cases of CRC will occur in 2040, with a concomitant 73.4% increase in deaths to over 1.6 million per annum (2). An ever-expanding body of evidence has identified several modifiable/lifestyle risk factors for CRC. Examples that have been associated with differential CRC risk include smoking, alcohol consumption, decreased dietary consumption of fiber, whole grains, vegetables, and fruits, living with obesity or being overweight, low physical activity, inflammatory bowel disease, and increased intake of red meat and processed meat (3–11). Red meat and processed meat have been labeled as “probably carcinogenic to humans” (group 2a carcinogen), and “carcinogenic to humans” (group 1 carcinogen), respectively, by the International Agency for Research on Cancer (IARC) due to the evidence vis-à-vis CRC (12). Different putative causal pathways have been proposed for the association between red meat and/or processed meat with CRC. Mainly, the presence of heme iron combined with long-term intestinal dysbiosis, in addition to the neoformation of mutagenic compounds (N-nitroso compounds, heterocyclic amines, polycyclic aromatic hydrocarbons) which have been reported to act both systemically, and locally at the mucosa of the colon and rectum (12–16); thus, potentially mediating the association between red meat and/or processed meat consumption and CRC risk.

The majority of CRC cases are considered sporadic, as over three-quarters of the patients do not have a family history (17). However, genome-wide association studies (GWAS) have informed a heritable basis for the disease, with over 200 single-nucleotide polymorphisms (SNPs) showing a marginally statistically significant association with the trait (18). Moreover, non-additive estimates of heritability such as gene-environment (GxE) interactions, have been reported to explain some variance in the heritability of CRC (19). GxE interactions refers to the scenario where the effect of a genetic variant differs across levels of the environmental exposure, or vice versa. That is, the susceptibility of a person to a given environmental exposure can be influenced by a genetic variant. Previously, most studies that evaluated GxE interactions in CRC were considered underpowered to detect these interactions (20). To address this, we recently published a genome-wide GxE interaction analysis that reported two significant novel interactions for red meat consumption: the rs4871179 SNP in chromosome (chr) 8 (downstream of *HAS2*) and the rs35352860 SNP in chr18 (*SMAD7* intronic region) (21). However, traditional candidate GxE analyses, which sequentially evaluate variants associated with the trait, prohibit the adequate identification of individuals at elevated CRC risk, and do not explain the underlying mechanisms of this highly polygenic carcinogenic process.

Polygenic risk scores (PRS) aggregate multiple SNPs reported through GWAS with a typically small effect to estimate heritability or genetic liability to a particular phenotype. In a classic PRS, a sum of the effect alleles that an individual carries, weighted by the effect sizes of the variants (e.g., GWAS summary statistics), is computed (22). This enables the output of a risk estimate at the individual level that could be leveraged to stratify patients and provide targeted screening, with the potential to guide precise CRC prevention (23). However, when modeled in relation to a phenotype, this canonical PRS approach assumes that every SNP that is an element of the PRS (i.e., *SNP_i_* ∈ *PRS*) exhibits the same estimate of association (i.e., *β_SNP_i__* =… = *β_SNP_n__*). Moreover, a loss of information ensues, as the PRS groups SNPs that are associated with the phenotype via dissimilar biological processes. To address this challenge, pathway-based PRS construct gene sets that aggregate risk alleles across *a priori* defined pathways via existing annotated databases (e.g., REACTOME, PANTHER Pathway, KEGG (24–26)). This strategy provides functionally informed risk scores that have enabled detection of putative disrupted biological pathways for various polygenic traits (e.g., Alzheimer’s disease, schizophrenia, coronary artery disease (27–29)) but has yet to be deployed for CRC risk.

In this study, we applied functional annotation data to identify subsets of GWAS variants that are components of biological pathways to construct pathway-based PRS (pPRS). Additionally, we evaluated multiplicative interactions between pPRS and red meat or processed meat intake and CRC risk in a large, pooled dataset to ascertain whether genes within specific pathways that exhibit a statistically significant interaction with the environmental factor could influence CRC risk.

## MATERIALS and METHODS

### Study participants

Details on study participants, genotyping information, and environmental exposure data have been previously published (30,31). In brief, 27 studies from the Genetics and Epidemiology of Colorectal Cancer Consortium (GECCO), the Colorectal Cancer Transdisciplinary Study (CORECT), and the Colon Cancer Family Registry (CCFR) were utilized. These studies have been previously described (21,32). The outcome/case status was defined as a positive diagnosis of invasive colorectal adenocarcinoma (International Classification of Diseases codes (ICD)153-154), and confirmed via medical records, pathology reports, or death certificate information. Nested case-control sets were assembled via risk-set sampling in cohort studies, and population-based controls were matched on age, sex, race, and enrollment date/trial group (in the SELECT trial) in case-control studies. Cases were further stratified by tumor location as follows: proximal colon (ICD-9 codes: 153.0, 153.1, 153.4, and 153.6), distal colon (ICD-9 codes: 153.2, 153.3, 153.7), and rectum (ICD-9 codes: 154.0, 154.1). Given the very small number participants of non-European descent across studies we only included individuals of European ancestry according to self-reported race and ethnicity and confirmation via genetic data. Exclusion criteria included: duplicate participants or cryptic relatedness, errors on genotyping and/or imputation, age outliers, diagnosis of advanced adenoma (only), and missing data for red meat and processed meat intake. All participants gave written informed consent, and studies were approved by their individual institutional review boards.

### Genotype data

Genotyping arrays utilized by each study are described in Supplementary Table S1. Quality control and genotyping metrics were conducted as previously described (30,31). Briefly, samples were excluded with the following criteria: 1) Single nucleotide polymorphisms (SNPs) with missing call rate >2-5%; 2) Departure from Hardy-Weinberg equilibrium (threshold = P<1×10-4); 3) Mismatch between genotyped and reported sex; 4) Discordant genotype calls with duplicate samples. The Haplotype Reference Consortium panel of 39.1 million variants was used to impute to the genotypes with the University of Michigan Imputation Server (33). We used the Binarydosage R package (https://cran.r-project.org/web/packages/BinaryDosage) to process and manage the imputed genotypes in our sample. Imputed SNPs with a minor allele frequency (MAF) <1% and/or imputation accuracy of R^2^ ≤ 0.8 were dropped. PLINK 1.9 was utilized to compute principal components on 30,000 randomly sampled SNPs with MAF >5% to control for population stratification (34). For use in polygenic risk score analyses, we leveraged data from 204 autosomal SNPs that have been previously reported to showcase a marginal genome-wide statistically significant association with CRC (P<5×10-8) (18).

### Environmental exposure data

In-person interviews, phone interviews, or structured self-administered questionnaires were utilized to collect environmental risk factors and sociodemographic variables as previously described (32). After quality-control checks in each study were conducted, common data elements (CDEs) were defined. Study-specific data elements were mapped to the CDEs through an iterative process with outreach to data contributors. Via SAS and a T-SQL script, all CDEs were pooled into a dataset that included common definitions, standardized permissible values, and standardized coding. The dataset was evaluated for quality assurance, with the distribution of the data within and between studies assessed for outlier observations. Food frequency questionnaires were utilized to compute total energy intake (kcal/day). Mean imputation was utilized for studies with partial missingness. In studies that did not report the variable (ASTERISK, DACHS, PHS, UKB) (Supplementary Table 1), the value was set to a constant value (zero). Body mass index (BMI) was computed as weight (kg)/height(m)^2^. The predefined World Health Organization BMI cut-off points were used to generate an ordinal categorical variable: underweight (<18.5 kg/m^2^), normal weight (18.5 to <25 kg/m^2^), overweight (≥25.0 to <30 kg/m^2^), and obese (≥30 kg/m^2^). Red meat intake was defined as consumption of beef, pork, and/or lamb, while processed meat included intake of bacon, sausages, luncheon/deli meats, and/or hot dogs. Both variables were expressed as servings per day (i.e., servings/day), with one serving equivalent to 70.9 grams or 2.5 ounces. Heterogeneity in the red meat/processed meat definition for individual studies caused overlap in some instances, with studies including processed meat under the red meat definition. We evaluated red meat and processed meat intake by constructing sex-study-specific quartiles, with the median of intake for each quartile utilized to define consumption values, fitted as continuous predictors in regression models.

### Pathway overrepresentation

From the 204 genome-wide significant autosomal SNPs reported by Fernandez-Rozadilla et. al (18), rsIDs were extracted in accordance with the genome reference consortium (GRC) human build 37 (GRCh37). These SNPs were then annotated using the Annotation Query (AnnoQ) platform, which provides integrated functional annotation supported by a database of ∼39 million pre-annotated human variants from the Haplotype Reference Consortium (35). Annotations to Ensembl and Reference sequence (RefSeq) genes were derived using inferences from Annotate Variation (ANNOVAR), SnpEff, and Ensembl Variant Effect Predictor (VEP) (36–40). SNPs residing in enhancer regions were linked to their target genes via the Predicted by Experimental Results: Enhancer-Gene Relationships Illustrated by a Nexus of Evidence (PEREGRINE), which leverages experimental data from chromatin interaction analysis by paired-end tag sequencing, expression quantitative trait locus, and Hi-C across 78 cell and tissue types (41). Duplicate gene identifiers were removed, and the resultant unique genes were uploaded into the Protein ANalysis THrough Evolutionary Relationships (PANTHER) classification system v.18.0. (42,43) which is built on 143 annotated complete genomes and assigns sequences to PANTHER Pathways (v3.6.7; 177 pathways and 3092 components) (25,43,44). The set of genes falling within each pathway were tested for overrepresentation relative to the PANTHER Pathway annotation sets using Fisher’s exact test, with false discovery rate correction applied for multiple testing (p < 0.05) (45). Each significantly overrepresented pathway was subsequently used for downstream analyses.

### Statistical analyses

Standard polygenic risk score (PRS) methods (22) were used to compute pathway-based polygenic risk scores (pPRS). SNP-specific weights were extracted after regressing CRC on the set of 204 variants in our sample with a logistic regression model that adjusted for study, sex, age, and three principal components of ancestry. A complete list of variants and weights can be found in Supplementary Table S2. The subset of weights corresponding to each of the overrepresented pathways were used to compute the pPRS. To identify potential interactions between pPRS and red meat/processed meat intake (pPRSxE), a 1 degree-of-freedom (1-df) test of gene-environment (GxE) interaction was utilized to determine statistical significance of the interaction term pPRSxE in multivariable logistic regression. All models were adjusted for study, age, sex, and the first three principal components to account for population structure. Simulations have shown that Type I error rates are conserved when the same sample is used to both generate PRS weights and to evaluate potential pPRSxE interactions (46). As variants can simultaneously participate in multiple pathways, pPRS subsets that included unique variants, and variants that showed overlap to other pathways were generated. To contrast the utility of deploying pPRS, we generated a PRS that included all 204 variants (overall PRS), and a PRS that contained the subset of these variants that were not overrepresented among PANTHER-defined pathways (no pathway PRS, n = 174). Potential effect modification of the association between red meat/processed meat intake and CRC risk by pPRS was evaluated by further stratifying the models by quartiles of the corresponding pPRS. All analyses were performed using the R programming language (v.4.2.1). All p-values are two-sided, with a value < 0.05 deemed statistically significant.

### Data availability

Data utilized to generate results in this study are available in the database of Genotypes and Phenotypes (dbGAP) (http://view.ncbi.nlm.nih.gov/dbgap-controlled).

## RESULTS

A total of 30,812 cases and 40,504 controls met the inclusion criteria. Of these, 138 had missing information on red meat intake (cases = 69, controls = 69) and 5,764 had missing processed meat intake information (cases = 2,634, controls = 3,130). Tumors were more commonly located in the proximal colon (35.2%), followed by the distal colon (30.4%), and the rectum (27.9%); 6.5% of the cases had a missing tumor location. Compared to controls, CRC cases, on average, had a higher BMI, were more likely to be male, and reported a higher total caloric intake. Cases had a higher mean intake of red meat (0.60 servings/day ± 0.46), and processed meat (0.35 servings/day ± 0.34) compared to controls (0.54 servings/day ± 0.44 & 0.28 ± 0.29, respectively, P-values < 0.001). Participant characteristics are described in detail in Supplementary Table S3.

### Pathway overrepresentation analysis and pathway-based polygenic risk scores

Pathway overrepresentation analysis revealed that 30 of the 204 SNPs that were *a priori* marginally associated with CRC were statistically significantly overrepresented across four biological pathways (p-values <0.001): 1) Gonadotropin-releasing hormone receptor pathway, n = 16 SNPs; 2) Presenilin-Alzheimer disease pathway, n = 8 SNPs; 3) Cadherin/WNT signaling pathway, n = 7 SNPs; and 4) Transforming Growth Factor – beta (TGF-β) signaling pathway, n = 14 SNPs (Supplementary Table S4). Some variants were simultaneously overrepresented in multiple pathways (Figure 1). The biggest overlap was reported between SNPs in the Gonadotropin-releasing hormone receptor pathway and TGF-β signaling pathway (n = 9). The rs35470271 SNP, which maps to chromosome 3 in the intergenic region of the *ZNF621* (Zinc Finger Protein 621), was the most commonly overrepresented SNP, with involvement in Gonadotropin-releasing hormone receptor pathway, presenilin-Alzheimer disease pathway, and Cadherin/WNT signaling pathway (Figure 1).

**Figure 1.**
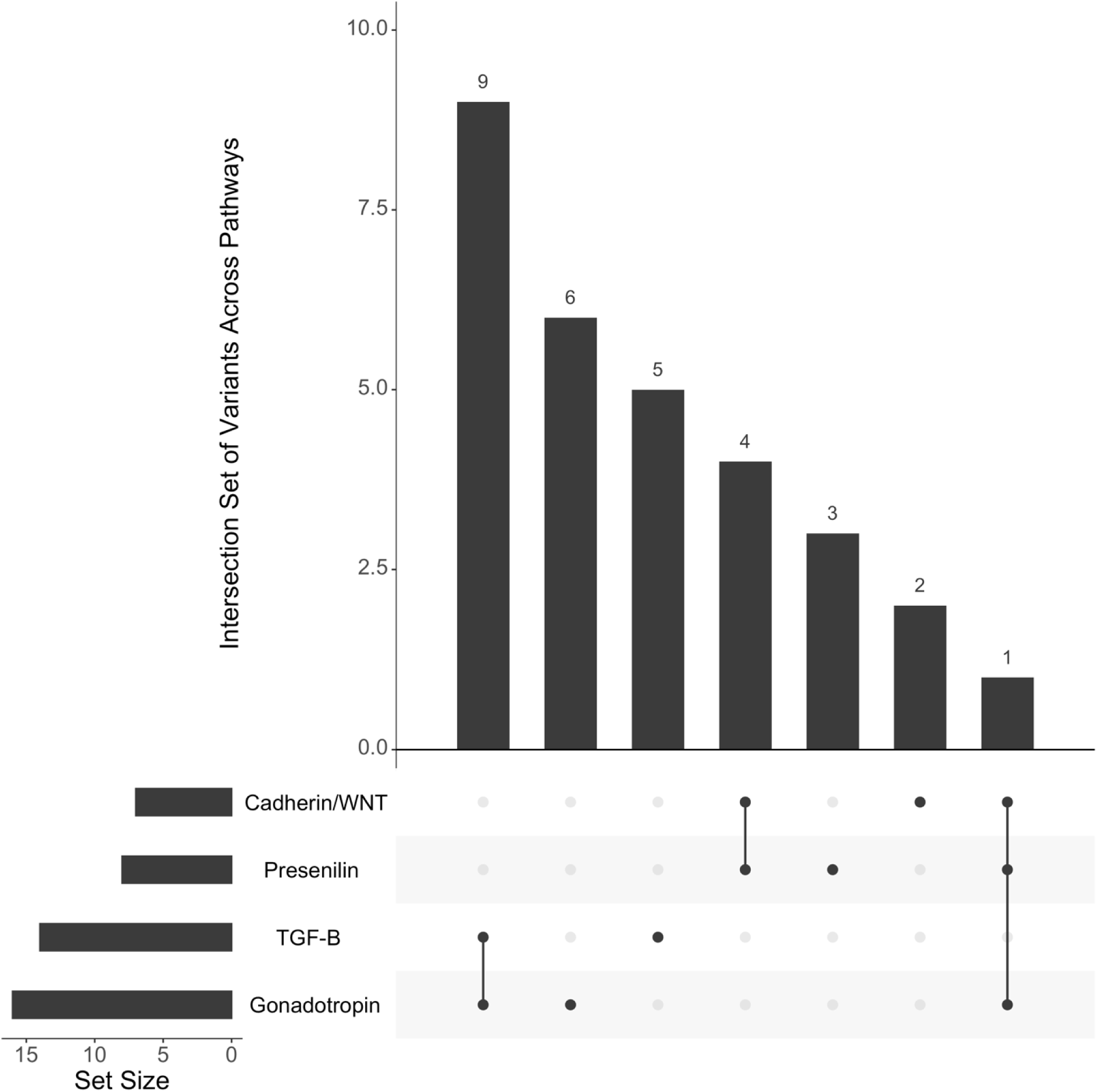
Overlap and set size of known colorectal cancer risk loci overrepresented across biological pathways. Cadherin/WNT, Cadherin/WNT signaling pathway; Presenilin, Presenilin-Alzheimer disease pathway; TGF-B, Transforming Growth Factor – beta signaling pathway; Gonadotropin, Gonadotropin-releasing hormone receptor pathway. Note: Set size corresponds to the number of single nucleotide polymorphisms contained in each pathway-based Polygenic Risk Score (pPRS).

Overrepresented and uniquely overrepresented SNPs in the four biological pathways were leveraged to construct pPRS. As expected, moderate to strong correlations were found between pPRS’s and their corresponding pPRS that contained only uniquely overrepresented SNPs (Figure 2). A strong correlation was found between the presenilin-Alzheimer disease-pPRS and Cadherin/WNT signaling-pPRS (r^2^ = 0.69), and between the TGF-β signaling-pPRS (TGF-β-PRS) and Gonadotropin-releasing hormone receptor-pPRS (r^2^ = 0.62) (Figure 2), driven by variants that overlapped across said pathways (Figure 1).

**Figure 2.**
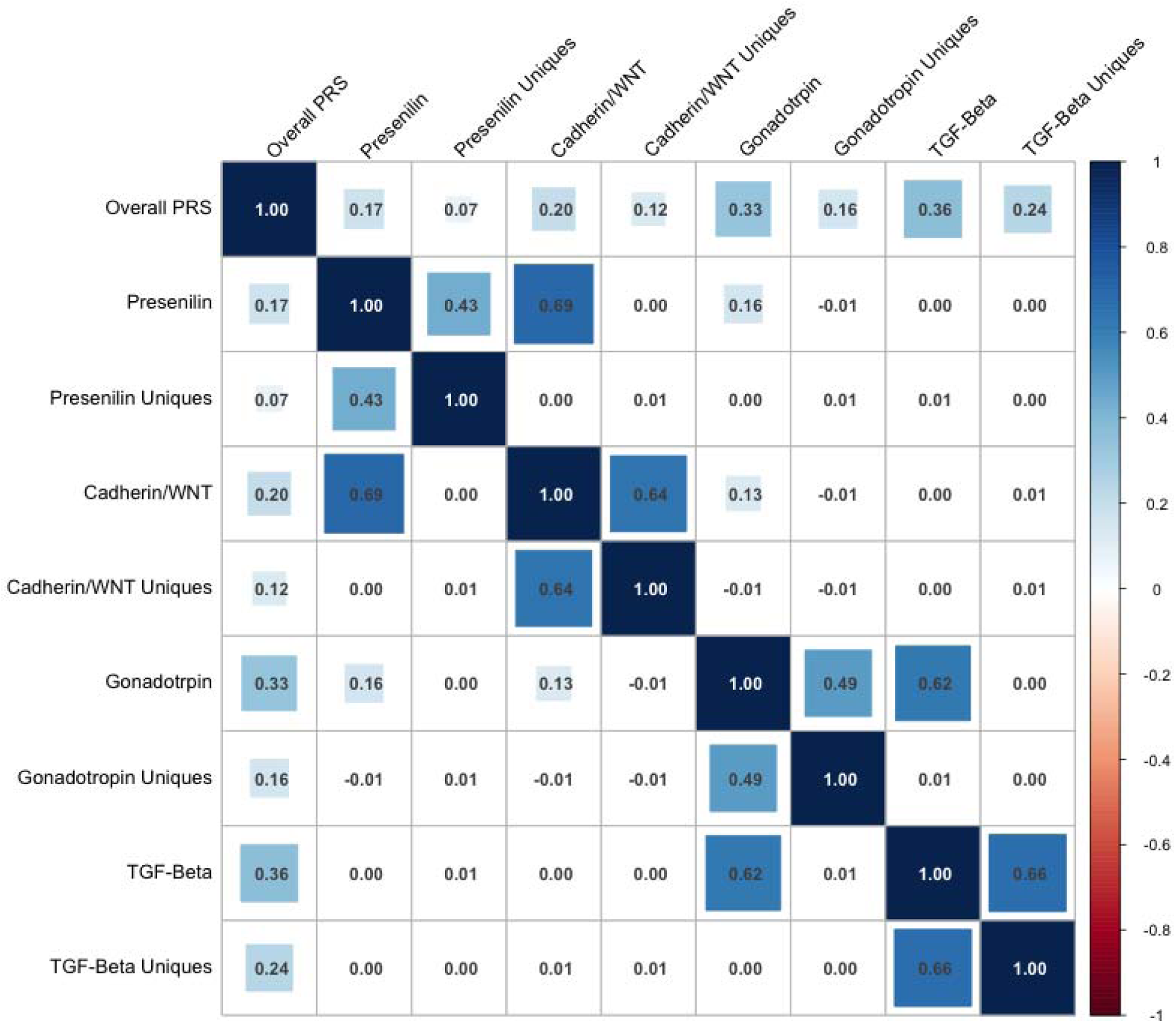
Correlogram for pathway-based polygenic risk scores (pPRS) constructed from known colorectal cancer risk loci overrepresented in biological pathways. Note: The numbers correspond to Pearson’s correlation coefficients. Overall PRS contains 204 SNPs that have been previously associated with colorectal cancer risk.

### Interaction between pPRS with red meat or processed meat intake in CRC risk

We observed a statistically significant interaction for the overall PRS with red meat intake (p-value = 0.0048). However, when we deployed the novel pPRS’s to evaluate the putative multiplicative interaction with meat intake (pPRSxE), the interaction was significant for the TGF-β-PRS with red meat intake (p-value = 0.0031), with no evidence of an interaction in the other three biological pathways. Moreover, the interaction remained significant when assessed in the TGF-β-PRS with only uniquely overrepresented variants for the TGF-β signaling pathway (p-value = 0.0007). No statistically significant interactions were identified for processed meat intake (Figure 3).

**Figure 3.**
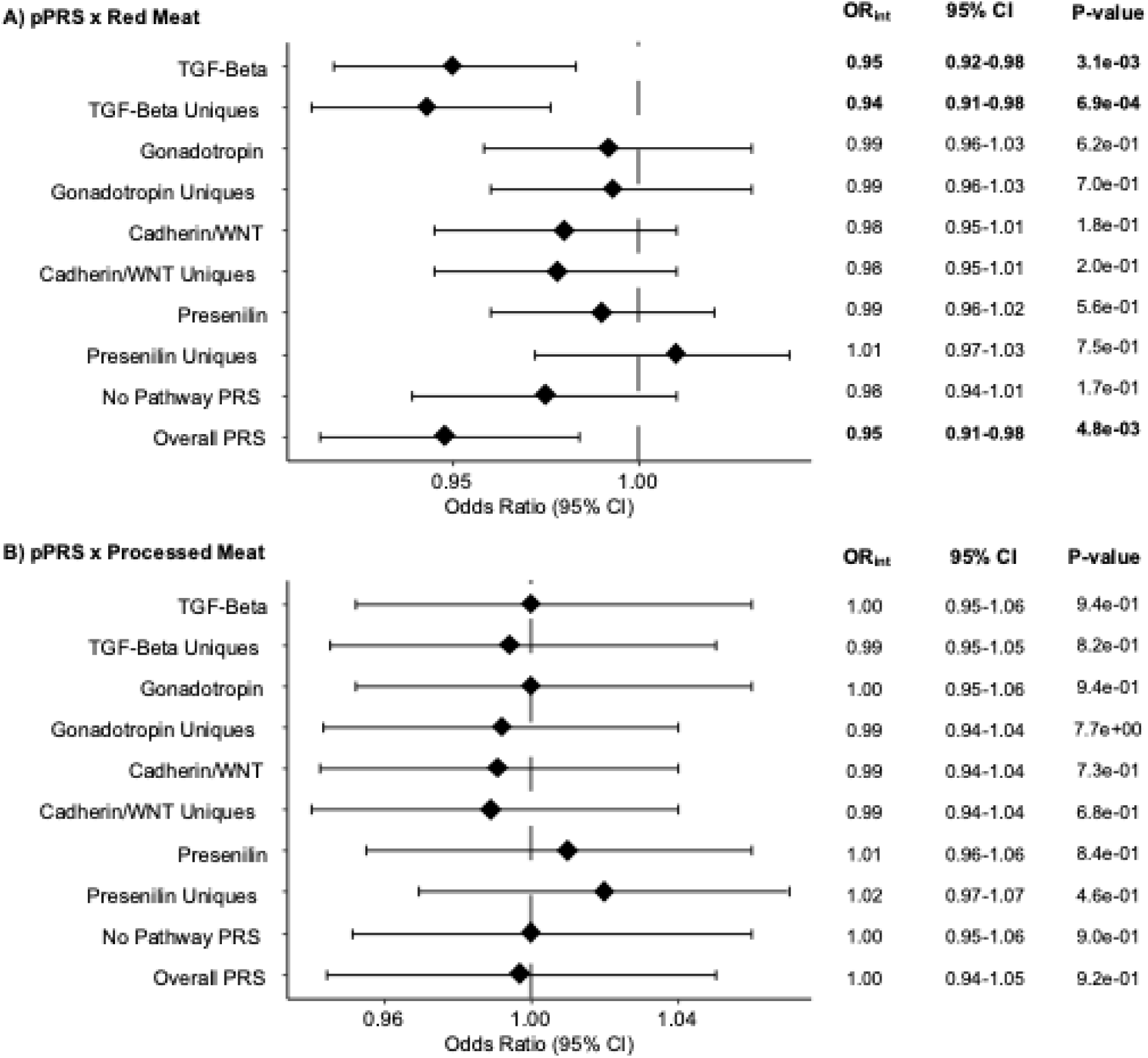
Odds Ratios and 95% Confidence Intervals for interactions between pathway-based Polygenic Risk Scores with **(A)** red meat, and **(B)** processed meat intake on colorectal cancer risk. pPRS, pathway-based Polygenic Risk Scores; OR, Odds Ratio; CI, Confidence Interval. Note: The No Pathway PRS contains variants that have previously shown a genome-wide significant association with colorectal cancer but were not overrepresented in biological pathways. For variants that are exclusively overrepresented in a single pathway, a second subset is generated (i.e., “Uniques”) and the appropriate pPRS computed. P-value corresponds to a 1-degree-of-freedom test of pPRSxE. All models were adjusted for study, age, sex, and the first three principal components.

When the pPRSxE interactions were examined in a multivariable multinomial logistic regression model with tumor location as the outcome variable (i.e., proximal colon, distal colon, and rectum), a statistically significant interaction was observed between the overall PRS, and the TGF-β-PRS and red meat intake in relation to CRC risk in the proximal colon and in the rectum (all p-values < 0.05) (Supplementary Table S5). In the TGF-β-PRS with only uniquely overrepresented variants for the TGF-β signaling pathway, a statistically significant interaction with red meat intake was identified for tumors in the proximal and distal colon (p-value = 0.0152 & p-value = 0.0245, respectively). Furthermore, we observed a statistically significant interaction between the TGF-β-PRS and processed meat intake for proximal colon adenocarcinomas (p-value = 0.0488) (Supplementary Table S5).

We stratified the association between red meat/processed meat intake in relation to CRC risk by quartiles of the distribution of the pPRS’s (Supplementary Table S6). After controlling for confounders, individuals in the ::S25^th^ percentile of the overall PRS stratum were 1.32 (95% CI = 1.21-1.44; p-value = 1.22×10-9), times as likely to be diagnosed with CRC associated with an increase red meat intake (median of servings/day from study-sex-specific quartiles), by contrast, individuals in the >75^th^ percentile of the overall PRS were only 1.17 times as likely (OR = 1.17; 95% CI = 1.07-1.28; p-value = 0.0005) (Table 1). Similarly, we observed that the association between red meat intake and CRC risk was stronger for individuals with lower genetic risk based on the TGF-β-PRS distribution. Specifically, individuals in the first (Q1), second (Q2), third (Q3), and fourth quartile (Q4) of the TGF-β-PRS had a 37%, 33%, 27%, and 17% higher CRC risk, respectively (Q1 OR = 1.37; 95% CI = 1.26-1.49; p-value = 1.20×10-13, Q2 OR = 1.33; 95% CI = 1.22-1.45; p-value = 4.39×10-11, Q3 OR = 1.27; 95% CI = 1.16-1.38; p-value = 5.45×10-08, Q4 OR = 1.17; 95% CI = 1.07-1.27; 4.41×10-04) (Table 1).

**Table 1.**
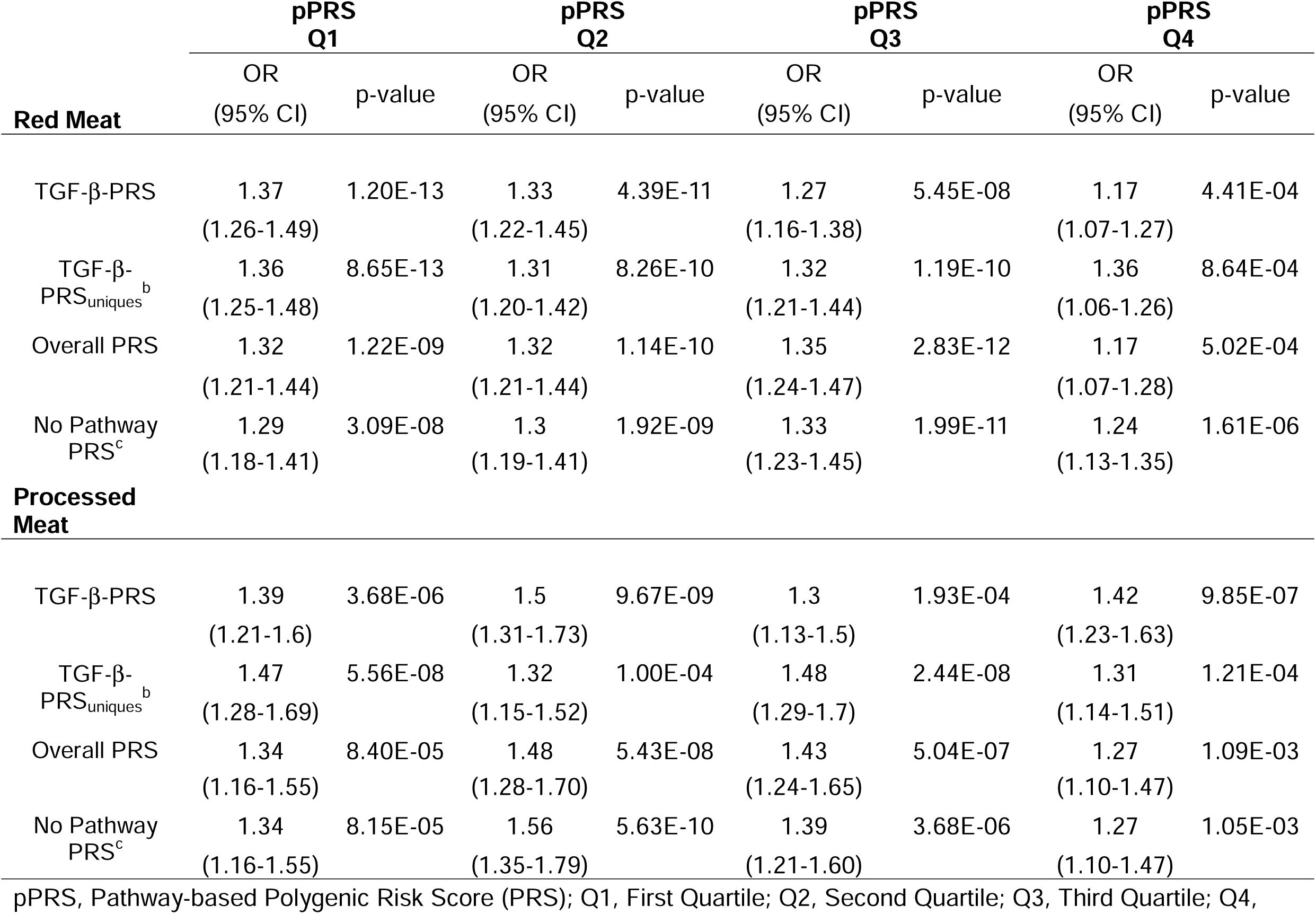
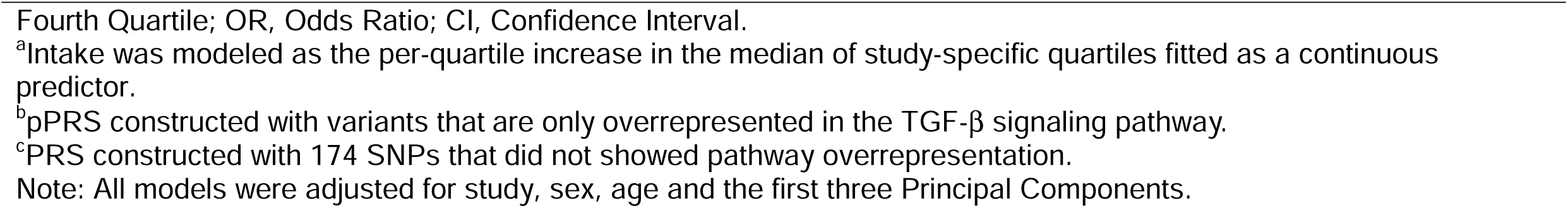
Association between red meat/processed meat intake^a^ with Colorectal Cancer risk stratified by quartiles of pPRS.

|  | pPRS<br>Q1 |  | pPRS<br>Q2 |  | pPRS<br>Q3 |  | pPRS<br>Q4 |  |
| --- | --- | --- | --- | --- | --- | --- | --- | --- |
|  | OR<br>(95% CI) | p-value | OR<br>(95% CI) | p-value | OR<br>(95% CI) | p-value | OR<br>(95% CI) | p-value |
| **Red Meat** |  |  |  |  |  |  |  |  |
| TGF-β-PRS | 1.37<br>(1.26-1.49) | 1.20E-13 | 1.33<br>(1.22-1.45) | 4.39E-11 | 1.27<br>(1.16-1.38) | 5.45E-08 | 1.17<br>(1.07-1.27) | 4.41E-04 |
| TGF-β-<br>PRS<sub>uniques</sub><sup>b</sup> | 1.36<br>(1.25-1.48) | 8.65E-13 | 1.31<br>(1.20-1.42) | 8.26E-10 | 1.32<br>(1.21-1.44) | 1.19E-10 | 1.36<br>(1.06-1.26) | 8.64E-04 |
| Overall PRS | 1.32<br>(1.21-1.44) | 1.22E-09 | 1.32<br>(1.21-1.44) | 1.14E-10 | 1.35<br>(1.24-1.47) | 2.83E-12 | 1.17<br>(1.07-1.28) | 5.02E-04 |
| No Pathway<br>PRS<sup>c</sup> | 1.29<br>(1.18-1.41) | 3.09E-08 | 1.3<br>(1.19-1.41) | 1.92E-09 | 1.33<br>(1.23-1.45) | 1.99E-11 | 1.24<br>(1.13-1.35) | 1.61E-06 |
| **Processed**<br>**Meat** |  |  |  |  |  |  |  |  |
| TGF-β-PRS | 1.39<br>(1.21-1.6) | 3.68E-06 | 1.5<br>(1.31-1.73) | 9.67E-09 | 1.3<br>(1.13-1.5) | 1.93E-04 | 1.42<br>(1.23-1.63) | 9.85E-07 |
| TGF-β-<br>PRS<sub>uniques</sub><sup>b</sup> | 1.47<br>(1.28-1.69) | 5.56E-08 | 1.32<br>(1.15-1.52) | 1.00E-04 | 1.48<br>(1.29-1.7) | 2.44E-08 | 1.31<br>(1.14-1.51) | 1.21E-04 |
| Overall PRS | 1.34<br>(1.16-1.55) | 8.40E-05 | 1.48<br>(1.28-1.70) | 5.43E-08 | 1.43<br>(1.24-1.65) | 5.04E-07 | 1.27<br>(1.10-1.47) | 1.09E-03 |
| No Pathway<br>PRS<sup>c</sup> | 1.34<br>(1.16-1.55) | 8.15E-05 | 1.56<br>(1.35-1.79) | 5.63E-10 | 1.39<br>(1.21-1.60) | 3.68E-06 | 1.27<br>(1.10-1.47) | 1.05E-03 |

pPRS, Pathway-based Polygenic Risk Score (PRS); Q1, First Quartile; Q2, Second Quartile; Q3, Third Quartile; Q4,
Fourth Quartile; OR, Odds Ratio; CI, Confidence Interval.
<sup>a</sup>Intake was modeled as the per-quartile increase in the median of study-specific quartiles fitted as a continuous predictor.
<sup>b</sup>pPRS constructed with variants that are only overrepresented in the TGF-β signaling pathway.
<sup>c</sup>PRS constructed with 174 SNPs that did not showed pathway overrepresentation.
Note: All models were adjusted for study, sex, age and the first three Principal Components.

### TGF-β-PRS and red meat intake interactions in CRC risk

We further interrogated the robustness of the statistically significant interaction between the TGF-β-PRS and red meat intake, upon further adjustment of the model with PRS’s that contained variants not overrepresented in the TGF-β signaling pathway, but that had been previously associated with CRC risk.

The interaction between the TGF-β-PRS and red meat intake remained significant after we included a fixed term with a PRS constructed from variants that showed no overlap with the TGF-β signaling pathway (n = 16) (p-value = 0.0040), with no change in the estimate of association for the interaction term (OR_int=_ 0.951; 95% CI = 0.919-0.984). Similarly, when we included PRS’s as model covariates for SNPs overrepresented in other pathways regardless of overlap with the TGF-β signaling pathway (n = 25), or with variants that are not overrepresented in the TGF-β signaling pathway, but are marginally associated with CRC risk (n = 190), the interaction between TGF-β-PRS and red meat intake was still significant (p -value = 0.0039 & p-value = 0.003, respectively) and the estimates of association remained consistent (OR_int_ = 0.951; 95% CI = 0.919-0.984 & OR_int_ = 0.949; 95% CI = 0.916-0.982). Furthermore, when we fitted a PRS constructed with the variants in the intersection between the TGF-β signaling-pPRS and the Gonadotropin-releasing hormone receptor-pPRS (i.e., overlap between both pathways, n = 9), no evidence of an interaction with red meat intake was observed (OR_int_ = 0.987; 95% CI = 0.954-1.021; p-value = 0.457), which suggests that the significance of the TGF-β-PRS interaction with red meat is driven by variants uniquely overrepresented in the TGF-β signaling pathway (Table 2). Sensitivity analyses confirmed that the TGF-β-PRS interaction with red meat was not confounded by smoking, BMI, consumption of alcohol (g/day), fiber (g/day), fruits (servings/day), vegetables (servings/day), or total caloric intake (kcal/day) (Supplementary Table S7).

**Table 2.** Multiplicative interactions between TGF-β-PRS and red meat intake in association with colorectal cancer risk.

| | TGF-β-PRS x Red Meat | | |
| --- | --- | --- | --- |
|  | OR<sub>int</sub> | 95% CI | P-interaction<sup>a</sup> |
| Model 1 | 0.950 | 0.918-0.983 | 0.0031 |
| Model 2 | 0.951 | 0.919-0.984 | 0.0040 |
| Model 3 | 0.951 | 0.919-0.984 | 0.0041 |
| Model 4 | 0.949 | 0.916-0.982 | 0.0030 |
| Model 5 | 0.987 | 0.954-1.021 | 0.4570 |

PRS, Polygenic Risk Score; OR, Odds Ratio; CI, Confidence Interval.
**Model 1:** Adjusted for study, sex, age and the first three Principal Components.
**Model 2:** Adjusted for covariates in Model 1, plus PRS that contains variants that were not overrepresented in the TGF-β pathway, but that showed statistically significant overrepresentation in other pathways (n = 16)
**Model 3:** Adjusted for covariates in Model 1, plus PRS that contains SNPs that do not exhibit overrepresentation for the TGF-β pathway but demonstrate overrepresentation in other pathways (n = 25).
**Model 4:** Adjusted for covariates in Model 1, plus a PRS constructed with all SNPs that do not showcase overrepresentation for the TGF-β pathway but have been marginally associated with the trait (n = 190).
**Model 5:** pPRS constructed with common variants between TGF-β , and Gonadotropin releasing hormone pathways (n = 9). Adjusted for covariates in Model 1.
<sup>a</sup> Computed with a 1 degree-of-freedom test.

When the variants that were uniquely overrepresented in the TGF-β signaling pathway were assessed separately, we found a statistically significant interaction with red meat consumption for rs2337113 which maps to chromosome 18 in an intronic region for the gene *SMAD7* (SMAD family member 7) (p-value = 0.0005), and for rs2208603, which maps to chromosome 6 in the intergenic region for *BMP5* (bone morphogenetic protein 5) (p-value = 0.036) (Table 3). Furthermore, we found limited evidence of a statistically significant interaction between the overall PRS and red meat intake on CRC risk when the five uniquely overrepresented SNPs in TGF-β signaling pathway were removed (OR_int_ = 0.96; 95% CI = 0.93-1.00; p-value = 0.0513).

**Table 3.** Gene-environment interactions (GxE) between single nucleotide polymorphisms (SNP) uniquely overrepresented in the TGF-β pathway^a^ and red meat intake in relation to colorectal cancer risk.

| SNP | Chr | BP Position | Effect | Gene | Ref | Alt | Alt Allele Freq (1000G EUR) | SNP x Red Meat |  |
| --- | --- | --- | --- | --- | --- | --- | --- | --- | --- |
|  |  |  |  |  |  |  |  | OR (95% CI) | P-interaction<sup>b</sup> |
| rs3809570 | 15 | 67000117 | Intronic/Upstream | *SMAD6* | C | A | 0.249 | 1.013<br>(0.958-1.071) | 0.6580 |
| rs2337113 | 18 | 46452327 | Intronic | *SMAD7* | A | G | 0.450 | 1.088<br>(1.037 -1.142) | 0.0005 |
| rs2208603 | 6 | 55577214 | Intergenic | *BMP5* | T | C | 0.708 | 0.946<br>(0.898-0.996) | 0.0360 |
| rs62404966 | 6 | 55712124 | Intronic | *BMP5* | C | T | 0.245 | 0.962<br>(0.909-1.019) | 0.1830 |
| rs6912214 | 6 | 55721302 | Intronic | *BMP5* | T | C | 0.458 | 1.019<br>(0.971-1.070) | 0.4430 |

SNP, single nucleotide polymorphism; Chr, chromosome; BP Position, base pair position based on GRCh37; OR, Odds Ratio; CI, Confidence Interval.
<sup>a</sup>SNPs that do not show overlap with any other pathway, except TGF-β .
<sup>b</sup>Computed with a 1 degree-of-freedom test.

## DISCUSSION

In this comprehensive analysis of more than 70,000 individuals, we report a statistically significant interaction between a PRS with red meat intake in relation to CRC risk. Furthermore, we constructed four pathway-based PRS’s which when deployed, suggested for the first time, that the significance of the PRS and red meat interaction was specifically driven by variants overrepresented in the TGF-β signaling pathway (Figure 3).

The TGF-β signaling pathway plays a biological role in cell proliferation, differentiation, migration, and apoptosis (47). Many of these roles are context-dependent and appear paradoxical. In benign tissues, TGF-β signaling inhibits epithelial growth and cell proliferation, while promoting differentiation, and apoptosis (47,48). Classically associated with epithelial cancers, TGF-β signaling acts as a tumor suppressor via promotion of cell cycle arrest and apoptosis during the tumor initiation phase (48). However, it facilitates tumor cell proliferation, epithelial-mesenchymal transition, fibrosis, inflammation, and cellular stemness via activation of the canonical and noncanonical TGF-β signaling pathways during tumor progression (49). Moreover, autocrine TGF-β signaling activation has the ability to change the architecture of the tumor microenvironment and suppress anti-tumor immune-mediated responses, enabling cell invasion, dissemination, and therapeutic resistance (50).

In CRC, an extensive body of work has reported on the involvement of this signaling pathway at multiple stages of the natural history of disease. Elevated TGF-β levels in plasma and primary tumor tissue correlate with higher tumor stage and recurrence (51,52), while its inhibition can prevent metastasis by inducing a potent cytotoxic T-cell response in murine models (53). Moreover, when the consensus molecular subgroup (CMS) classification for CRC was proposed, a distinctive transcriptional signature for TGF-β signaling, which conveyed the worst relapse-free survival of all 4 CMS subgroups, was reported (i.e., CMS-4) (54,55). The potential for an interaction between the TGF-β signaling pathway with red meat intake in relation to CRC risk remains understudied. Nevertheless, it has been suggested that oxysterols and aldehydes, formed through the oxidation of saturated fat in red meat, may upregulate TGF-β1 expression in macrophages and fibroblasts in the tumor vicinity (56). This sustained exposure to elevated concentrations of TGF-β1 in the colonic mucosa could facilitate clonal proliferation. However, this theory is not specific to red meat intake, as other sources of saturated fat exist (57). Additionally, we found no evidence to suggest that the TGF-β-PRS x red meat interaction was confounded by body mass index or total caloric intake (Supplementary Table S7). Therefore, it is unlikely that our findings are solely driven by increased saturated fat intake.

It is possible, that the underlying mechanism by which the TGF-β signaling pathway predisposes to an increased CRC risk rests in downstream components. Moreover, elements of this pathway, including GREM1, SMAD3, SMAD7, SMAD9, BMP2, BMP4, and RHPN2 have been linked to CRC etiology (47). Thus, in our study, we propose that polymorphisms targeting key elements of the canonical TGF-β signaling pathway interact with red meat intake to affect CRC risk (Table 3).

The rs2337113 SNP, which maps to the *SMAD7* gene, codes for an intracellular protein that is considered a negative regulator of TGF-β1 (58). Located in a region (long arm) of chromosome 18 that is commonly deleted in CRC (59,60), multiple *SMAD7* polymorphisms have been associated with CRC risk in Asian and European populations (61–64). In addition, deletion of *SMAD7* was associated with a protective dose effect in overall survival and disease-free survival in 264 tumor biopsies of colorectal cancer patients, while amplification was associated with poor prognosis (65). The involvement of the rs2337113 variant in CRC risk is supported by the work of Tian et al. (66). Furthermore, this consortium previously published a genome-wide statistically significant interaction between another *SMAD7* polymorphism (rs35352860, chr18:48927384, C>T) with red meat intake in relation to CRC risk (21), which showcases a strong correlation (ρ = 0.642; 95% CI = 0.638-0.646; p-value = 2.2E-16) with the rs2337113 SNP (A>G). In our previous publication, we hypothesized that inhibition of the liver-derived peptide hepcidin via overexpression of rs35352860 could prevent the internalization of the transmembrane receptor ferroportin with a concomitant increase in iron output into the bloodstream, which in turn, could facilitate free radical formation and a proinflammatory, procardiogenic state. This aligns with the work by Stolfi et al., which demonstrated that SMAD7 silencing inhibited the growth of CRC cell lines both in vitro and in vivo after transplantation into immunodeficient mice. Additionally, a significant increase in SMAD7 expression was observed in 14 matched pairs of CRC and adjacent tissues (67). Thus, our finding of an interaction between rs2337113 and red meat, provides additional evidence to support the potential involvement of *SMAD7* in CRC risk.

The other significant interaction between red meat and individual SNPs in the TGF-β signaling pathway was for a variant in the 6p12.1 region (rs2208603) which resides in the intergenic region of the gene *BMP5.* This gene codes for a cytokine member of the bone morphogenetic proteins (BMPs)/growth differentiation factors; a protein class within the TGF-β superfamily (68). BMP5 binds to the activin receptor-like kinase (ALK) −2 and −6, which controls binding to BMP type II and I receptors to induce SMAD-dependent, or SMAD-independent, intracellular signaling (69). This signaling is inhibited by SMAD7, as it preferentially interacts with type I receptors (70). Albeit BMPs have been reported to exhibit cell-dependent and context-dependent inhibitory capabilities (71–73). In CRC, deep sequencing of sporadic CRC samples identified loss of *BMP5* as an early and CRC-specific event, while high expression correlated with longer survival. Moreover, knockdown of *BMP5* in the SW480 cell line (adenocarcinoma of the colon, Dukes’ type B) promoted proliferation. Thus, suggesting its relevance as a tumor suppressor gene (74). However, mutational events are uncommon; only <4% prevalence of non-silent somatic mutations in CRC was found in TCGA samples (75). Further research is required to evaluate the potential interaction between *BMP5* and red meat intake in relation to CRC. However, recent work from Xiao et al., highlighted the role of BMP5 as a regulator of hepcidin and systemic iron homeostasis. In BMP5-inactive murine models, exposure to an iron-poor or high-iron diet decreased hepcidin levels in young mice. This transcriptional inactivation of hepcidin was exacerbated when concomitant BMP6 inactivation ensued, leading to hepatic and extrahepatic iron overload. Notably, the iron overload became more severe in animals exposed to a high-iron diet (76). This further supports our *a priori* mentioned hypothesis in which functional inhibition of hepcidin could foster a pro-carcinogenic state, increasing CRC risk.

Although previous literature has published on the association between the Gonadotropin-releasing hormone pathway, presenilin-Alzheimer disease pathway, and Cadherin/WNT signaling pathway with CRC risk (77–79), we found no evidence of a statistically significant interaction with red meat or processed meat intake on CRC risk (Figure 3). However, the observed overlap of SNPs overrepresented across multiple pathways suggests crosstalk and co-regulation between biological pathways (Figure 1). Prime examples are the multilevel processes jointly regulated by the TGF-β signaling pathway and Cadherin/WNT signaling. In conjunction, these pathways control extracellular gradients of morphogens during embryonic development, regulate target gene expression at the nucleus, and protein-protein interactions in the cytoplasm. Moreover, TGF-β and WNT synergistically promote tumorigenesis (80). Thus, further research is required to elucidate the potential antagonistic or synergistic effects of multilevel, multifactor interactions across these biological pathways in relation to red meat and/or processed meat intake for CRC.

Our study has several strengths. We leveraged harmonized pooled data that underwent systematic quality control, enabling the consideration of tumor localization, study design, and potential confounders of the exposure and outcome in the putative causal pathway. Additionally, the implementation of the novel pathway-based PRS x E approach by Gauderman et al. (46) allowed us not only to detect an interaction between SNPs that previously showed a marginal association with CRC with red meat intake in a single score, but also to identify the variants driving this signal and underpin the pathway leading to increased CRC risk. This approach supports the construction of constrained PRSs with variants selected according to a priori established criteria (e.g., pathway overrepresentation) (Figure 2). This is in line with a previous study, that reported no linear correlation between the predictive accuracy of PRSs and the number of SNPs in the score (81). A key strength of our approach lies in its comprehensive nature: we integrate annotations from SnpEff, ANNOVAR, and VEP, drawing from both Ensembl and RefSeq databases to enhance the robustness of SNP-to-gene mapping. To further account for regulatory variants, we incorporated PEREGRINE-based enhancer–gene link annotations, enabling the capture of potential functional effects from non-coding SNPs. Although we recognize that incorporating study-specific eQTL or chromatin interaction data would offer an even finer resolution of regulatory effects, our multi-tool strategy substantially reduces annotation discrepancies and improves the accuracy of linking non-coding variants to their target genes. Nonetheless, we also recognize some limitations. Alternative tools and databases, such as Reactome (reactome.org) or Gene Ontology (geneontology.org), offer different pathway definitions that may not fully overlap with those in PANTHER, albeit no gold standard exists. However, when Reactome was utilized to annotate variants, the signaling by TGF-β family members pathway showed evidence of activation (FDR p-value = 0.04). In addition, different methods to explore novel GxE interactions and subsequently conduct pathway analyses have been reported (82). Nevertheless, our approach, which leverages *a priori* reported variants that have been marginally associated with the trait of interest, allowed us to propose a more parsimonious set of putative causal SNPs that, in conjunction with red meat intake, may be acting through similar biological processes to influence CRC risk. Predominantly, questionnaire data was utilized to construct the red and processed meat intake variables and given that many of the included studies have a case-control design, participants were asked to report on intake 1-2 years before study selection. Thus, we cannot rule out the potential for misclassification bias of the exposure and/or recall bias. In cohort studies, exposures were assessed at the time of blood draw or buccal collection at study-specific times (83). Also, we utilize study-specific quartiles to evaluate meat intake, which does not account for absolute differences. However, this approach is a valid method in pooled analysis of nutritional exposures (84). Furthermore, total caloric intake (kcal/day) was not reported in four studies and set to zero (ASTERISK, DACHS, PHS, UKB). Albeit sensitivity analyses confirmed that, adjustment with a fixed indicator for study in the interaction models, prevented those four studies to contribute information for estimating the energy effect. However, they did contribute to the estimation of other effects and the pPRSxE interaction term. In addition, the interaction between TGF-β-PRS and red meat was not confounded by total caloric intake (kcal/day). Finally, our findings may not be generalizable to other racial and ethnic populations besides those of European ancestry, which contributed to the individual studies.

## CONCLUSION

In summary, this pathway-based interaction analysis proposes, for the first time, that a statistical interaction between SNPs overrepresented in the TGF-β signaling pathway and red meat intake impacts CRC risk. This finding provides a potential mechanistic explanation linking red meat consumption and CRC risk and highlights the importance of dietary guidance and personalized prevention strategies.

## AKNOWLEDGMENTS

ASTERISK: We are very grateful to Dr. Bruno Buecher without whom this project would not have existed. We also thank all those who agreed to participate in this study, including the patients and the healthy control participants, as well as all the physicians, technicians, and students. CCFR: The Colon CFR graciously thanks the generous contributions of their study participants, dedication of study staff, and the financial support from the U.S. National Cancer Institute, without which this important registry would not exist. The authors would like to thank the study participants and staff of the Seattle Colon Cancer Family Registry and the Hormones and Colon Cancer study (CORE Studies). CLUE II: We thank the participants of Clue II and appreciate the continued efforts of the staff at the Johns Hopkins George W. Comstock Center for Public Health Research and Prevention in the conduct of the Clue II Cohort Study. Cancer data was provided by the Maryland Cancer Registry, Center for Cancer Prevention and Control, Maryland Department of Health, with funding from the State of Maryland and the Maryland Cigarette Restitution Fund. The collection and availability of cancer registry data is also supported by the Cooperative Agreement NU58DP006333, funded by the Centers for Disease Control and Prevention. Its contents are solely the responsibility of the authors and do not necessarily represent the official views of the Centers for Disease Control and Prevention or the Department of Health and Human Services. CPS-II: The authors express sincere appreciation to all Cancer Prevention Study-II participants, and to each member of the study and biospecimen management group. The authors would like to acknowledge the contribution to this study from central cancer registries supported through the Centers for Disease Control and Prevention’s National Program of Cancer Registries and cancer registries supported by the National Cancer Institute’s Surveillance Epidemiology and End Results Program. The authors assume full responsibility for all analyses and interpretation of results. The views expressed here are those of the authors and do not necessarily represent the American Cancer Society or the American Cancer Society – Cancer Action Network. DACHS: We thank all participants and cooperating clinicians, and everyone who provided excellent technical assistance. EDRN: We acknowledge all contributors to the development of the resource at University of Pittsburgh School of Medicine, Department of Gastroenterology, Department of Pathology, Hepatology and Nutrition and Biomedical Informatics. EPIC: Where authors are identified as personnel of the International Agency for Research on Cancer/World Health Organization, the authors alone are responsible for the views expressed in this article and they do not necessarily represent the decisions, policy, or views of the International Agency for Research on Cancer/World Health Organization. Harvard cohorts: The study protocol was approved by the institutional review boards of the Brigham and Women’s Hospital and Harvard T.H. Chan School of Public Health, and those of participating registries as required. We acknowledge Channing Division of Network Medicine, Department of Medicine, Brigham, and Women’s Hospital as home of the NHS. The authors would like to acknowledge the contribution to this study from central cancer registries supported through the Centers for Disease Control and Prevention’s National Program of Cancer Registries (NPCR) and/or the National Cancer Institute’s Surveillance, Epidemiology, and End Results (SEER) Program. Central registries may also be supported by state agencies, universities, and cancer centers. Participating central cancer registries include the following: Alabama, Alaska, Arizona, Arkansas, California, Colorado, Connecticut, Delaware, Florida, Georgia, Hawaii, Idaho, Indiana, Iowa, Kentucky, Louisiana, Massachusetts, Maine, Maryland, Michigan, Mississippi, Montana, Nebraska, Nevada, New Hampshire, New Jersey, New Mexico, New York, North Carolina, North Dakota, Ohio, Oklahoma, Oregon, Pennsylvania, Puerto Rico, Rhode Island, Seattle SEER Registry, South Carolina, Tennessee, Texas, Utah, Virginia, West Virginia, Wyoming. The authors assume full responsibility for analyses and interpretation of these data. Kentucky: We would like to acknowledge the staff at the Kentucky Cancer Registry. LCCS: We acknowledge the contributions of Jennifer Barrett, Robin Waxman, Gillian Smith and Emma Northwood in conducting this study. NCCCS I & II: We would like to thank the study participants, and the NC Colorectal Cancer Study staff. PLCO: The authors thank the PLCO Cancer Screening Trial screening center investigators and the staff from Information Management Services Inc and Westat Inc. Most importantly, we thank the study participants for their contributions that made this study possible. Cancer incidence data have been provided by the District of Columbia Cancer Registry, Georgia Cancer Registry, Hawaii Cancer Registry, Minnesota Cancer Surveillance System, Missouri Cancer Registry, Nevada Central Cancer Registry, Pennsylvania Cancer Registry, Texas Cancer Registry, Virginia Cancer Registry, and Wisconsin Cancer Reporting System. All are supported in part by funds from the Center for Disease Control and Prevention, National Program for Central Registries, local states or by the National Cancer Institute, Surveillance, Epidemiology, and End Results program. The results reported here, and the conclusions derived are the sole responsibility of the authors. SELECT: We thank the research and clinical staff at the sites that participated on SELECT study, without whom the trial would not have been successful. We are also grateful to the 35,533 dedicated men who participated in SELECT. WHI: The authors thank the WHI investigators and staff for their dedication, and the study participants for making the program possible. A full listing of WHI investigators can be found at: http://www.whi.org/researchers/Documents%20%20Write%20a%20Paper/WHI%20Investigator%20Short%20List.pdf

## FUNDING

MCS and JSM received support from awards U54CA233465 and U2CCA252971 from the National Cancer Institute. Genetics and Epidemiology of Colorectal Cancer Consortium (GECCO): National Cancer Institute, National Institutes of Health, U.S. Department of Health and Human Services (U01 CA137088, R01 CA059045, U01 CA164930, R01 CA201407). This research was funded in part through the NIH/NCI Cancer Center Support Grant P30 CA015704 and P30CA014089. Scientific Computing Infrastructure at Fred Hutch funded by ORIP grant S10OD028685. ASTERISK: a Hospital Clinical Research Program (PHRC-BRD09/C) from the University Hospital Center of Nantes (CHU de Nantes) and supported by the Regional Council of Pays de la Loire, the Groupement des Entreprises Françaises dans la Lutte contre le Cancer (GEFLUC), the Association Anne de Bretagne Génétique and the Ligue Régionale Contre le Cancer (LRCC). The ATBC Study is supported by the Intramural Research Program of the U.S. National Cancer Institute, National Institutes of Health, Department of Health and Human Services. CLUE II funding was from the National Cancer Institute (U01 CA086308, Early Detection Research Network; P30 CA006973), National Institute on Aging (U01 AG018033), and the American Institute for Cancer Research. The content of this publication does not necessarily reflect the views or policies of the Department of Health and Human Services, nor does mention of trade names, commercial products, or organizations imply endorsement by the US government. The Colon Cancer Family Registry (CCFR, www.coloncfr.org) is supported in part by funding from the National Cancer Institute (NCI), National Institutes of Health (NIH) (award U01 CA167551). Support for case ascertainment was provided in part from the Surveillance, Epidemiology, and End Results (SEER) Program and the following U.S. state cancer registries: AZ, CO, MN, NC, NH; and by the Victoria Cancer Registry (Australia) and Ontario Cancer Registry (Canada). The CCFR Set-1 (Illumina 1M/1M-Duo) and Set-2 (Illumina Omni1-Quad) scans were supported by NIH awards U01 CA122839 and R01 CA143237 (to GC). The CCFR Set-3 (Affymetrix Axiom CORECT Set array) was supported by NIH award U19 CA148107 and R01 CA81488 (to SBG). The CCFR Set-4 (Illumina OncoArray 600K SNP array) was supported by NIH award U19 CA148107 (to SBG) and by the Center for Inherited Disease Research (CIDR), which is funded by the NIH to the Johns Hopkins University, contract number HHSN268201200008I. The content of this manuscript does not necessarily reflect the views or policies of the NCI, NIH or any of the collaborating centers in the Colon Cancer Family Registry (CCFR), nor does mention of trade names, commercial products, or organizations imply endorsement by the US Government, any cancer registry, or the CCFR. COLO2&3: National Institutes of Health (R01 CA060987). CPS-II: The American Cancer Society funds the creation, maintenance, and updating of the Cancer Prevention Study-II (CPS-II) cohort. The study protocol was approved by the institutional review boards of Emory University, and those of participating registries as required. CRCGEN: Colorectal Cancer Genetics & Genomics, Spanish study was supported by Instituto de Salud Carlos III, co-funded by FEDER funds –a way to build Europe– (grants PI14-613 and PI09-1286), Agency for Management of University and Research Grants (AGAUR) of the Catalan Government (grant 2017SGR723), Junta de Castilla y León (grant LE22A10-2), the Spanish Association Against Cancer (AECC) Scientific Foundation grant GCTRA18022MORE and the Consortium for Biomedical Research in Epidemiology and Public Health (CIBERESP), action Genrisk. Sample collection of this work was supported by the Xarxa de Bancs de Tumors de Catalunya sponsored by Pla Director d’Oncología de Catalunya (XBTC), Plataforma Biobancos PT13/0010/0013 and ICOBIOBANC, sponsored by the Catalan Institute of Oncology. We thank CERCA Programme, Generalitat de Catalunya for institutional support. DACHS: This work was supported by the German Research Council (BR 1704/6-1, BR 1704/6-3, BR 1704/6-4, CH 117/1-1, HO 5117/2-1, HE 5998/2-1, KL 2354/3-1, RO 2270/8-1 and BR 1704/17-1), the Interdisciplinary Research Program of the National Center for Tumor Diseases (NCT), Germany, and the German Federal Ministry of Education and Research (01KH0404, 01ER0814, 01ER0815, 01ER1505A, 01ER1505B and 01KD2104A). DALS: National Institutes of Health (R01 CA048998 to M. L. Slattery). EPIC: The coordination of EPIC is financially supported by International Agency for Research on Cancer (IARC) and also by the Department of Epidemiology and Biostatistics, School of Public Health, Imperial College London which has additional infrastructure support provided by the NIHR Imperial Biomedical Research Centre (BRC). The national cohorts are supported by: Danish Cancer Society (Denmark); Ligue Contre le Cancer, Institut Gustave Roussy, Mutuelle Générale de l’Education Nationale, Institut National de la Santé et de la Recherche Médicale (INSERM) (France); German Cancer Aid, German Cancer Research Center (DKFZ), German Institute of Human Nutrition Potsdam-Rehbruecke (DIfE), Federal Ministry of Education and Research (BMBF) (Germany); Associazione Italiana per la Ricerca sul Cancro-AIRC-Italy, Compagnia di SanPaolo and National Research Council (Italy); Dutch Ministry of Public Health, Welfare and Sports (VWS), Netherlands Cancer Registry (NKR), LK Research Funds, Dutch Prevention Funds, Dutch ZON (Zorg Onderzoek Nederland), World Cancer Research Fund (WCRF), Statistics Netherlands (The Netherlands); Health Research Fund (FIS) - Instituto de Salud Carlos III (ISCIII), Regional Governments of Andalucía, Asturias, Basque Country, Murcia and Navarra, and the Catalan Institute of Oncology - ICO (Spain); Swedish Cancer Society, Swedish Research Council and and Region Skåne and Region Västerbotten (Sweden); Cancer Research UK (14136 to EPIC-Norfolk; C8221/A29017 to EPIC-Oxford), Medical Research Council (1000143 to EPIC-Norfolk; MR/M012190/1 to EPIC-Oxford). (United Kingdom). Harvard cohorts: HPFS is supported by the National Institutes of Health (P01 CA055075, UM1 CA167552, U01 CA167552, R01 CA137178, R01 CA151993, and R35 CA197735), NHS by the National Institutes of Health (P01 CA087969, UM1 CA186107, R01 CA137178, R01 CA151993, and R35 CA197735), and PHS by the National Institutes of Health (R01 CA042182). Kentucky: This work was supported by the following grant support: Clinical Investigator Award from Damon Runyon Cancer Research Foundation (CI-8); NCI R01CA136726. LCCS: The Leeds Colorectal Cancer Study was funded by the Food Standards Agency and Cancer Research UK Programme Award (C588/A19167). MCCS cohort recruitment was funded by VicHealth and Cancer Council Victoria. Melbourne Collaborative Cohort Study (MCCS) cohort recruitment was funded by VicHealth and Cancer Council Victoria. The MCCS was further augmented by Australian National Health and Medical Research Council grants 209057, 396414 and 1074383 and by infrastructure provided by Cancer Council Victoria. Cases were ascertained through the Victorian Cancer Registry. MEC: National Institutes of Health (R37 CA054281, P01 CA033619, and R01 CA063464). MECC: This work was supported by the National Institutes of Health, U.S. Department of Health and Human Services (R01 CA081488, R01 CA197350, U19 CA148107, R01 CA242218, and a generous gift from Daniel and Maryann Fong. NCCCS I & II: We acknowledge funding support for this project from the National Institutes of Health, R01 CA066635 and P30 DK034987. PLCO: Intramural Research Program of the Division of Cancer Epidemiology and Genetics and supported by contracts from the Division of Cancer Prevention, National Cancer Institute, NIH, DHHS. Funding was provided by National Institutes of Health (NIH), Genes, Environment and Health Initiative (GEI) Z01 CP 010200, NIH U01 HG004446, and NIH GEI U01 HG 004438. SELECT: Research reported in this publication was supported in part by the National Cancer Institute of the National Institutes of Health under Award Numbers U10 CA037429 (CD Blanke), and UM1 CA182883 (CM Tangen/IM Thompson). The content is solely the responsibility of the authors and does not necessarily represent the official views of the National Institutes of Health.

Swedish Mammography Cohort and Cohort of Swedish Men: This work is supported by the Swedish Research Council /Infrastructure grant, the Swedish Cancer Foundation, and the Karolinska Institutés Distinguished Professor Award to Alicja Wolk. VITAL: National Institutes of Health (K05 CA154337). WHI: The WHI program is funded by the National Heart, Lung, and Blood Institute, National Institutes of Health, U.S. Department of Health and Human Services through contracts HHSN268201600018C, HHSN268201600001C, HHSN268201600002C, HHSN268201600003C, and HHSN268201600004C. The authors thank the WHI investigators and staff for their dedication, and the study participants for making the program possible. A full listing of WHI investigators can be found at: https://www-whi-org.s3.us-west-2.amazonaws.com/wp-content/uploads/WHI-Investigator-Long-List.pdf

## Notes

**Conflicts of interest:** C.M.U. has as cancer center director oversight over research funded by several pharmaceutical companies but has not received funding directly herself. U.P was a consultant with AbbVie and her family is holding individual stocks for the following companies: Amazon, Boeing Company, BioNTech, BYD Company Limited, Crowdstrike Holdings Inc, CureVac, Google/Alphabet, Microsoft Corp, MicroStrategy Inc, NVIDIA Corp, Stellantis. M.G. receives research funding from Janssen and Sunbird Bio and consulting fees from Nerviano Medical Sciences, all unrelated to this work. C.E.T is an epidemiology contract with Pfizer, unrelated to the present study.

### Competing Interest Statement

C.M.U. has as cancer center director oversight over research funded by several pharmaceutical companies but has not received funding directly herself.
U.P was a consultant with AbbVie and her family is holding individual stocks for the following companies: Amazon, Boeing Company, BioNTech, BYD Company Limited, Crowdstrike Holdings Inc, CureVac, Google/Alphabet, Microsoft Corp, MicroStrategy Inc, NVIDIA Corp, Stellantis.
M.G. receives research funding from Janssen and Sunbird Bio and consulting fees from Nerviano Medical Sciences, all unrelated to this work.
C.E.T is an epidemiology contract with Pfizer, unrelated to the present study.

### Author Declarations

The study used ONLY available human data that was already published in the respective studies.

### Summary of Updates

Author string has been revised and article formatted for better readability.

